# Remote auditory task performance, cognition, and APOE4 across two dementia-risk cohorts

**DOI:** 10.64898/2026.11.19.26368469

**Authors:** Chris Happs, Arina Toma, Ivan Koychev, Paresh Malhotra, John T. O’Brien, Craig W. Ritchie, Graciela Muniz-Terrera, Meher Lad

## Abstract

**Background:** Difficulty hearing speech in noise is associated with cognitive ageing and dementia risk. It is unclear whether performance on remote auditory tasks reflects the presence of the apolipoprotein E (APOE) ε4 allele, the strongest common genetic risk factor for Alzheimer’s disease, or broader cognitive and ageing processes.

**Methods:** We analysed two independent cohorts: the Great Minds research register of older adults and the mid-life PREVENT Dementia programme. Both completed the same remote auditory battery: digits-in-noise, speech-in-babble, auditory working-memory precision for tone frequency and for amplitude-modulation rate, and the Goldsmiths Musical Sophistication Index. Cognitive outcomes were paired-associate learning in Great Minds, and the Four Mountains Task, Supermarket Task and selected COGNITO scores in PREVENT. Associations were tested with covariate-adjusted robust regression and false-discovery-rate (FDR) correction.

**Results:** The analytic samples comprised 164 Great Minds and 90 PREVENT participants (mean age 70.6 vs 56.8 years). APOE ε4 carrier status was not associated with any auditory measure in either cohort (all FDR-corrected p>0.70). In Great Minds, better speech-in-babble performance and better amplitude-modulation-rate working-memory precision were each associated with better paired-associate learning (both FDR-corrected p=0.019). In PREVENT, no auditory measure was associated with spatial-memory or COGNITO scores after age and sex adjustment and FDR correction.

**Conclusions:** Remote auditory task performance was associated with memory in the older cohort but not with APOE ε4 carrier status in either cohort. Longitudinal studies are needed to determine whether remote auditory testing predicts cognitive change.

## Introduction

Hearing difficulties in noisy environments are among the earliest and most common sensory complaints in older adults. Beyond communication challenges, such impairments have been linked to accelerated cognitive decline and an increased risk of dementia [1]. The mechanisms underlying this hearing–cognition relationship remain debated: they may be primarily peripheral, reflecting cochlear damage, or central, reflecting shared neurocognitive resources [2]. A growing body of work suggests that speech-in-noise perception depends not only on hearing sensitivity but also on cognitive mechanisms such as attention and auditory working memory [3], and these mechanisms are affected in Alzheimer’s disease dementia [4].

The APOE ε4 allele is the major common genetic risk factor for late-onset Alzheimer’s disease (AD) and is associated with earlier amyloid accumulation and hippocampal dysfunction [5]. Inheriting one copy of the ε4 allele increases AD risk roughly 3–4 times, and two copies 8–15 times [6]. Cognitively unimpaired ε4 carriers also perform worse than non-carriers on measures of episodic memory, executive function and global cognition [7]. Because auditory processing and memory share temporal-lobe and hippocampal networks, APOE ε4 status may influence auditory performance alongside cognition. Evidence that APOE ε4 is associated with remote central-auditory task performance, however, remains limited.

Previous laboratory work has shown that short-term memory for sound features, such as frequency and amplitude-modulation rate, contributes to speech-in-noise perception and correlates with cognitive screening performance. Lad et al. [3] showed that auditory memory for temporal fluctuations predicted both speech-in-noise performance and Addenbrooke’s Cognitive Examination scores, suggesting shared mechanisms between auditory memory and broader cognition. Web-based versions of these paradigms have since been shown to give results comparable to laboratory testing [8], making it feasible to study auditory–genetic– cognitive relationships at scale and remotely.

Examining this question in more than one cohort, from mid-life to late life, is informative because remote auditory phenotyping may behave differently across the dementia-risk continuum. We therefore analysed two independent datasets. The Great Minds register includes older adults with paired-associate learning (PALTEA) scores and APOE genotype. The PREVENT Dementia programme is a mid-life cohort designed to identify risk factors and the earliest biological changes associated with Alzheimer’s disease; available measures included APOE ε4 status, Four Mountains Task (4MT) and Supermarket Task (SMT) spatial-memory scores, and selected COGNITO computerised cognitive measures. The contrast in age and risk context allows the genetic and cognitive associations to be tested at two distinct life stages.

First, in the older Great Minds cohort, we examined APOE ε4–auditory associations and auditory predictors of PALTEA. Second, in the mid-life PREVENT cohort, we repeated the APOE ε4– auditory analysis and examined associations between auditory measures and spatial memory (4MT, SMT) and computerised cognition (COGNITO). We hypothesised that (1) APOE ε4 carriers would show poorer auditory performance than non-carriers, and (2) auditory measures, particularly speech-in-noise tasks, would be associated with cognitive performance.

## Materials and methods

### Study design

This was a cross-sectional secondary analysis of remote auditory task performance in two independent cohorts with genetic and cognitive data. Analyses were performed separately within each cohort because the samples differ strongly in age and cognitive outcomes, and results are presented side by side.

### Ethics statement

This study was approved by the Newcastle University Faculty of Medical Sciences Ethics Committee (Reference: 2682/41403). All participants gave written (electronic) informed consent before taking part. This was a retrospective analysis of existing cohort data. Great Minds data were accessed for research purposes through the DPUK Data Portal from 16 September 2025, and PREVENT data from 7 May 2026. All data were pseudonymised before access, and the authors had no access to information that could identify individual participants during or after data collection.

### Participants and data sources

Great Minds is part of Dementias Platform UK (DPUK) and is a register of middle-aged and older adults who have consented to be approached for brain-health research and to complete remote cognitive, genetic and lifestyle assessments [9]. For this analysis it was accessed through the DPUK Data Portal virtual machine under a data access agreement. In total, 201 participants had APOE genotype and remote auditory data. After excluding 37 participants who reported using loudspeakers, the analytic sample comprised 164 participants; after additionally excluding APOE ε2 carriers, the primary genetic sample comprised 147 participants (107 ε4 non-carriers, 40 carriers). Available variables included age, sex, education, APOE genotype, PALTEA, the auditory measures and headphone type.

PREVENT is a multi-site prospective cohort study in the UK and Ireland that recruited cognitively healthy adults aged 40–59 years who undergo cognitive testing, APOE genotyping and dementia-risk assessment [10,11]. Around half of the cohort have a parental history of dementia [11]. The speaker-excluded PREVENT sample comprised 90 participants from UK sites; headphone type was not recorded for 8 of these, who were retained because no loudspeaker use was reported. Of the 90, 71 had APOE genotype data and were not ε2 carriers (42 non-carriers, 29 carriers), and 63 of these had complete data for the APOE models.

### Auditory measures

Both cohorts completed five previously validated remote auditory measures [4,8]: digits-in-noise (DIN), speech-in-babble (SIB), auditory working-memory precision for tone frequency (WM-F) and for amplitude-modulation rate (WM-AM), and the Goldsmiths Musical Sophistication Index (GMSI) total score.

In the DIN task, participants listened to three digits presented in speech-shaped noise generated from the long-term average speech spectrum (80–10,000 Hz). Two practice trials were given at a signal-to-noise ratio (SNR) of 10 dB. An adaptive 1-up, 1-down procedure reduced the SNR after a correct response and increased it after an error. The starting SNR was 0 dB and the step size decreased from 5 to 2 dB after 3 reversals, then to 0.5 dB after 3 further reversals. The run ended after 10 reversals, and the SNRs at the last 5 reversals were averaged to give the DIN threshold (dB SNR); lower thresholds indicate better performance.

The SIB task used sentences presented in 16-talker babble [12,13]. Target sentences had the form <name> <verb> <number> <adjective> <noun> (e.g., “Alan gives four pretty flowers”), and participants selected the correct word from five columns of options. The SIB threshold was obtained with the same adaptive procedure and starting parameters as the DIN task.

Auditory working memory was tested with non-speech stimuli [14]. A 1-second pure tone or amplitude-modulated white-noise stimulus was presented, after which participants reproduced it by clicking along a horizontal on-screen scale, each click playing a sound at that position, and advanced when satisfied with the match. Tone frequencies were drawn from a uniform distribution between 440 and 880 Hz and amplitude-modulation rates from 5 to 20 Hz. Hanning windows were applied to all sounds. The task comprised 32 interleaved frequency and amplitude-modulation trials with a break after 16 trials. The SD of matching errors was estimated by fitting a Gaussian function, and its inverse (precision) was used for analysis, giving WM-F and WM-AM; higher precision indicates better performance [15].

Participants completed the 38-item GMSI self-report questionnaire [16]; the total score was the sum of the Active Engagement, Perceptual Abilities, Musical Training, Singing Abilities and Emotions subscales.

### Cognitive outcomes

In Great Minds, the cognitive outcome was PALTEA (Paired Associates Learning Total Errors Adjusted) from the CANTAB battery, expressed as a z-score relative to the CANTAB normative database with adjustment for age and sex [17]. The z-score is signed so that higher values indicate better performance (fewer errors than expected). In PREVENT, the spatial-memory outcomes were 4MT (15 items) and SMT (12 items) total scores, treated as missing when fewer than 14 4MT items or 11 SMT items were completed. Six COGNITO outcomes were available: name–face binding, immediate and delayed name recall, dual-task visual attention, semantic fluency and description recall. Higher 4MT, SMT and COGNITO scores indicate better performance.

### Device and headphone quality control

Participants received an emailed link to an online test session and were asked to complete all tests in one sitting, in a quiet room, on a tablet or laptop, using in-ear or over-ear headphones. Because headphone use could not be verified, participants reported at the end of the session whether they had used in-ear headphones, over-ear headphones or loudspeakers. Loudspeaker sessions were excluded from all analyses because of the risk of poor-quality stimulus delivery.

### Genetic variables

APOE genotype was coded as ε4 carrier status (at least one ε4 allele; ε3/ε4 and ε4/ε4 combined) and ε2 carrier status. Primary genetic analyses excluded ε2 carriers because this allele may be protective and could confound the comparison [18].

### Statistical analysis

Analyses were performed in Python (version 3.11) using pandas [19], SciPy [20] and statsmodels [21]. Continuous variables are reported as mean (SD) and categorical variables as n (%). Cohort differences were tested with Welch’s t-tests for continuous variables and chi-square tests for categorical variables.

Auditory measures were z-standardised within cohort. For all models, auditory scores were oriented so that higher values indicate better performance: DIN and SIB thresholds were sign-reversed (lower SNR thresholds became higher scores), whereas WM-F, WM-AM and GMSI are already scored so that higher is better. All models were ordinary least-squares regressions with heteroscedasticity-consistent (HC3) standard errors, fitted to complete cases, and all tests were two-sided.

Two families of models were fitted. (1) APOE ε4 models: each z-standardised auditory measure was regressed on ε4 carrier status (non-carriers as reference). In Great Minds these models adjusted for age, sex, education and PALTEA, as specified in the preregistered full model; in PREVENT they adjusted for age and sex. The coefficient is the difference between carriers and non-carriers in SD units of the auditory measure. (2) Auditory–cognition models: each cognitive outcome was regressed on one z-standardised auditory measure at a time, adjusted for age and sex, and additionally for education in Great Minds. The coefficient is the change in the outcome (PALTEA z-score units, or raw task score points in PREVENT) per 1 SD better auditory performance. In PREVENT, 4MT and SMT scores from visit 2 were used as primary outcomes; COGNITO outcomes were available from visit 1; as these were the closest to the time of the auditory assessments. Unadjusted Pearson correlations between auditory measures and 4MT/SMT were also calculated in PREVENT.

Benjamini–Hochberg false-discovery-rate (FDR) correction was applied across the five auditory measures within each cohort for the APOE ε4 models, and across the five auditory predictors within each cognitive outcome for the auditory–cognition models and correlations. Statistical significance was defined as an FDR-corrected p-value (q) < 0.05. Uncorrected p-values and q-values are reported. Full model results are given in S1 Table.

## Results

### Cohort characteristics

The speaker-excluded analytic samples comprised 164 Great Minds and 90 PREVENT participants. The cohorts differed substantially in age (Great Minds 70.6 (7.0) years vs PREVENT 56.8 (6.1) years; p<0.001) but were similar in the proportion of women (59% vs 62%; p=0.747), degree-level education (68% vs 73%; p=0.429) and in-ear headphone use (43% vs 45%; p=0.892). The APOE ε4 carrier rate was numerically higher in PREVENT (38% vs 27%; p=0.115). PREVENT participants performed better on all four auditory tasks and reported higher musical sophistication, consistent with their younger age (Table 1, Fig 1).

**Fig 1.**
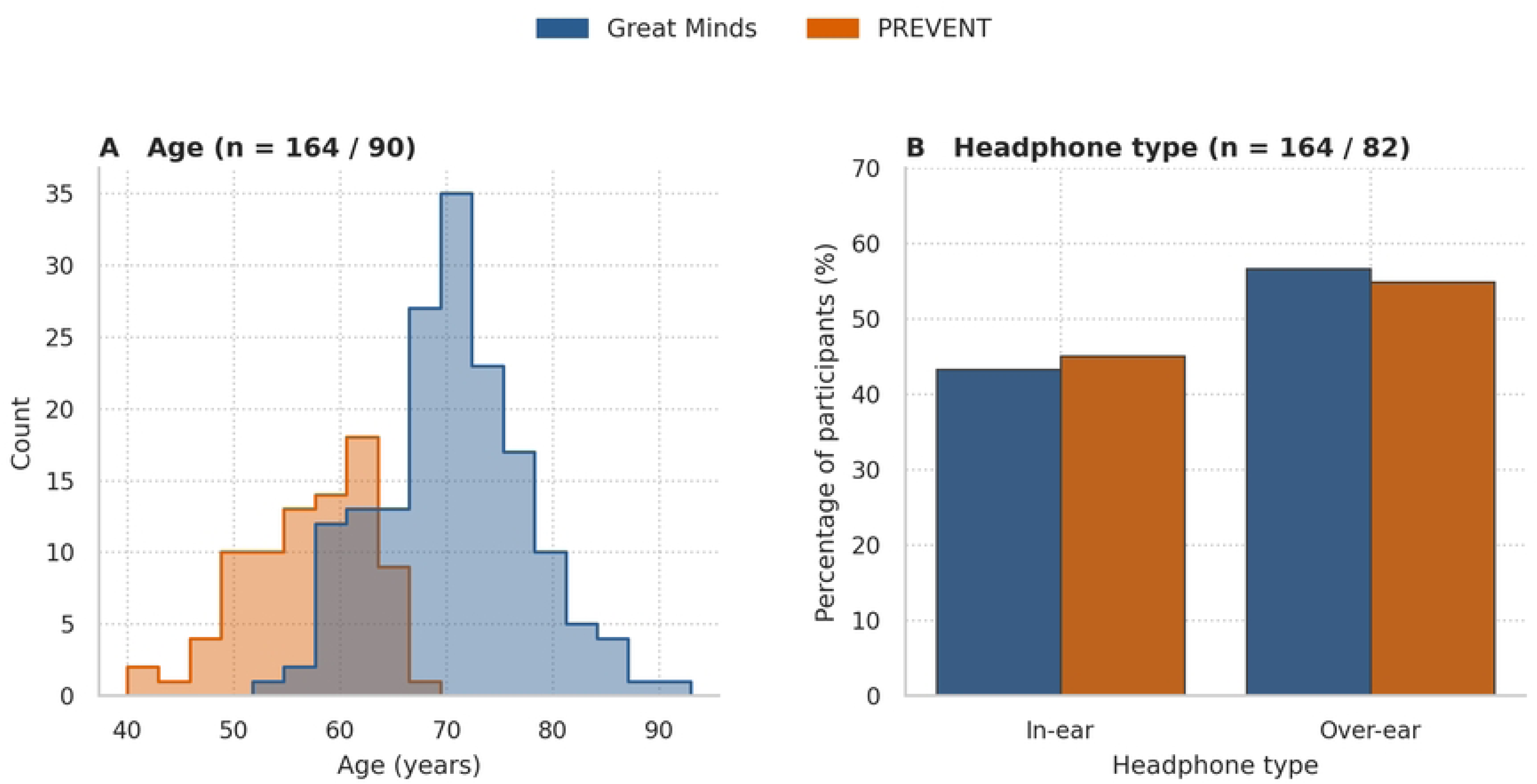
Cohort profile and online testing context. (A) Age distributions of the speaker-excluded Great Minds (n=164) and PREVENT (n=90) samples. (B) Percentage of participants using in-ear or over-ear headphones among those with headphone type recorded (Great Minds n=164, PREVENT n=82).

**Table 1.** Cohort characteristics and remote auditory performance.

|  | **Great Minds** | **PREVENT** | **p** |
|---|---|---|---|
| N | 164 | 90 | – |
| Age, years | 70.6 (7.0) | 56.8 (6.1) | <0.001 |
| Female sex | 97 (59%) | 51 (62%) | 0.747 |
| Degree or above | 111 (68%) | 61 (73%) | 0.429 |
| APOE ε4 carrier | 45 (27%) | 32 (38%) | 0.115 |
| In-ear headphones | 71 (43%) | 37 (45%) | 0.892 |
| DIN threshold, dB SNR | −3.6 (6.6) | −5.8 (3.7) | 0.001 |
| SIB threshold, dB SNR | 1.1 (3.7) | −1.2 (4.5) | <0.001 |
| WM-F precision | 0.27 (0.17) | 0.36 (0.26) | 0.007 |
| WM-AM precision | 0.08 (0.03) | 0.10 (0.04) | 0.002 |
| GMSI total | 141.3 (20.6) | 148.5 (17.1) | 0.004 |

Values are mean (SD) or n (%). Percentages are of participants with available data; PREVENT denominators were 82–84 (sex and headphone type, n=82). APOE ε4 carrier counts include ε2/ε4 carriers. p-values from Welch’s t-tests (continuous) or chi-square tests (categorical). Lower DIN and SIB thresholds and higher WM-F, WM-AM and GMSI scores indicate better performance. DIN, digits-in-noise; SIB, speech-in-babble; WM-F, working-memory precision for tone frequency; WM-AM, working-memory precision for amplitude-modulation rate; GMSI, Goldsmiths Musical Sophistication Index; SNR, signal-to-noise ratio.

### APOE ε4 and auditory performance

APOE ε4 carrier status was not associated with any auditory measure in either cohort after adjustment and FDR correction (Table 2, Fig 2). The largest estimates were for WM-F in both cohorts (Great Minds β=−0.25, 95% CI −0.62 to 0.12, p=0.181; PREVENT β=−0.29, 95% CI −0.82 to 0.23, p=0.274) and for SIB in Great Minds (β=−0.20, 95% CI −0.57 to 0.17, p=0.292). Across all speech-in-noise, working-memory and musical-sophistication measures, all uncorrected p-values were >0.15 and all q-values >0.70, and every 95% confidence interval included zero.

**Fig 2.**
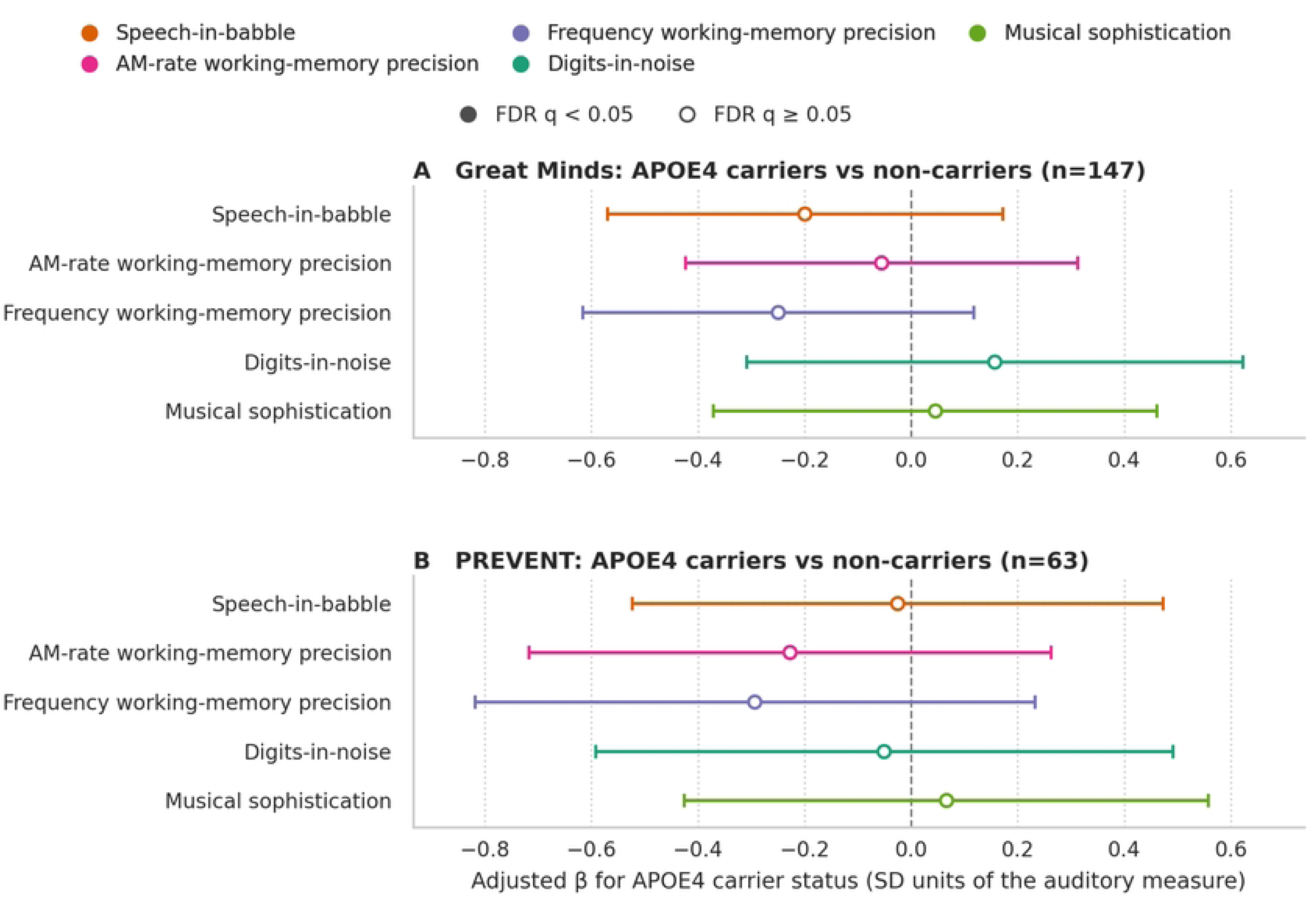
APOE ε4 carrier status and auditory performance in each cohort. Points show adjusted regression coefficients (β) for ε4 carriers versus non-carriers, in SD units of each auditory measure (higher = better performance), with 95% confidence intervals. (A) Great Minds (n=145–147), adjusted for age, sex, education and PALTEA. (B) PREVENT (n=63), adjusted for age and sex. Ordinary least-squares regression with HC3 robust standard errors. Open circles indicate FDR q ≥ 0.05 (no estimate reached q < 0.05).

**Table 2.** APOE ε4 carrier status and remote auditory performance.

| **Auditory measure** | **Cohort** | **n** | **β** | **95% CI** | **p** | **q (FDR)** |
|---|---|---|---|---|---|---|
| SIB | Great Minds | 147 | −0.20 | −0.57 to 0.17 | 0.292 | 0.730 |
| SIB | PREVENT | 63 | −0.03 | −0.52 to 0.47 | 0.919 | 0.919 |
| WM-AM | Great Minds | 147 | −0.06 | −0.42 to 0.31 | 0.767 | 0.833 |
| WM-AM | PREVENT | 63 | −0.23 | −0.72 to 0.26 | 0.363 | 0.907 |
| WM-F | Great Minds | 147 | −0.25 | −0.62 to 0.12 | 0.181 | 0.730 |
| WM-F | PREVENT | 63 | −0.29 | −0.82 to 0.23 | 0.274 | 0.907 |
| DIN | Great Minds | 146 | 0.16 | −0.31 to 0.62 | 0.510 | 0.833 |
| DIN | PREVENT | 63 | −0.05 | −0.59 to 0.49 | 0.854 | 0.919 |
| GMSI | Great Minds | 145 | 0.04 | −0.37 to 0.46 | 0.833 | 0.833 |
| GMSI | PREVENT | 63 | 0.07 | −0.43 to 0.56 | 0.794 | 0.919 |

β is the adjusted difference between APOE ε4 carriers and non-carriers (reference) in SD units of the auditory measure, oriented so that higher values indicate better performance. ε2 carriers were excluded. Great Minds models were adjusted for age, sex, education and PALTEA (preregistered full model); PREVENT models for age and sex. Ordinary least-squares regression with HC3 robust standard errors. q, Benjamini–Hochberg FDR-corrected p-value across the five auditory measures within each cohort. Abbreviations as in Table 1.

### Auditory predictors of cognition

In Great Minds, better SIB performance (β=0.20, 95% CI 0.06 to 0.33, p=0.006, q=0.019) and better WM-AM precision (β=0.20, 95% CI 0.05 to 0.35, p=0.008, q=0.019) were each associated with better PALTEA scores (Table 3, Fig 3A). DIN, WM-F and GMSI were not associated with PALTEA.

**Fig 3.**
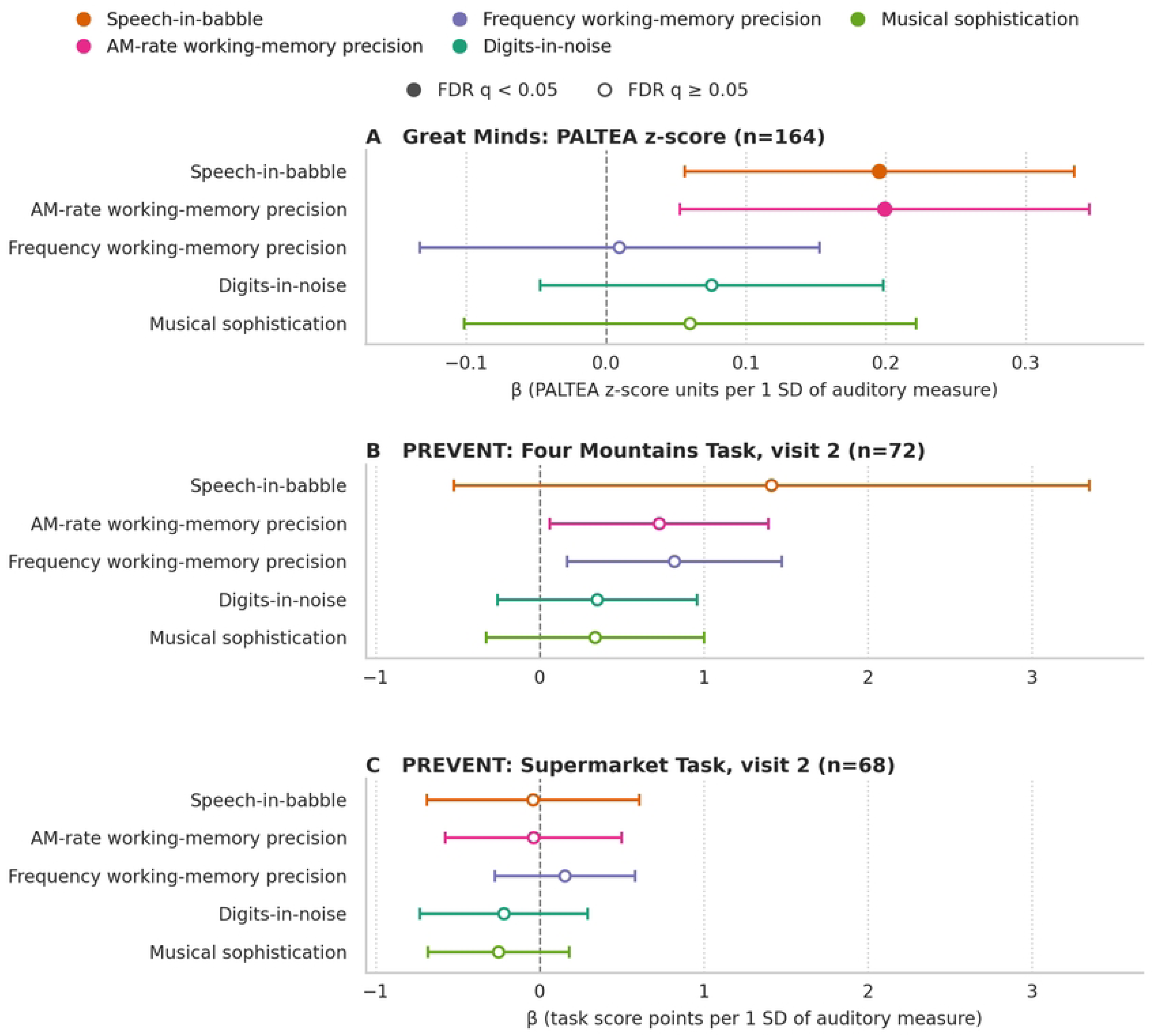
Auditory predictors of PALTEA, Four Mountains and Supermarket scores. Points show regression coefficients (β) per 1 SD better auditory performance, with 95% confidence intervals. (A) Great Minds PALTEA z-score (higher = better), adjusted for age, sex and education. (B) PREVENT Four Mountains Task and (C) Supermarket Task total scores at visit 2 (task score points), adjusted for age and sex. Ordinary least-squares regression with HC3 robust standard errors. Filled circles indicate FDR q < 0.05 within outcome.

**Table 3.** Auditory predictors of PALTEA, Four Mountains and Supermarket scores.

| **Cohort, outcome** | **Auditory predictor** | **n** | **β** | **95% CI** | **p** | **q (FDR)** |
|---|---|---|---|---|---|---|
| Great Minds, PALTEA | SIB | 164 | 0.20 | 0.06 to 0.33 | 0.006 | 0.019 |
|  | WM-AM | 164 | 0.20 | 0.05 to 0.35 | 0.008 | 0.019 |
|  | WM-F | 164 | 0.01 | −0.13 to 0.15 | 0.896 | 0.896 |
|  | DIN | 163 | 0.08 | −0.05 to 0.20 | 0.230 | 0.383 |
|  | GMSI | 162 | 0.06 | −0.10 to 0.22 | 0.467 | 0.584 |
| PREVENT, 4MT (visit 2) | SIB | 72 | 1.41 | −0.53 to 3.35 | 0.153 | 0.255 |
|  | WM-AM | 72 | 0.73 | 0.06 to 1.39 | 0.032 | 0.081 |
|  | WM-F | 72 | 0.82 | 0.16 to 1.47 | 0.014 | 0.071 |
|  | DIN | 72 | 0.35 | −0.26 to 0.96 | 0.259 | 0.320 |
|  | GMSI | 72 | 0.34 | −0.33 to 1.00 | 0.320 | 0.320 |
| PREVENT, SMT (visit 2) | SIB | 68 | −0.04 | −0.69 to 0.61 | 0.898 | 0.898 |
|  | WM-AM | 68 | −0.04 | −0.58 to 0.50 | 0.887 | 0.898 |
|  | WM-F | 68 | 0.15 | −0.27 to 0.58 | 0.484 | 0.807 |
|  | DIN | 68 | −0.22 | −0.73 to 0.29 | 0.399 | 0.807 |
|  | GMSI | 68 | −0.25 | −0.68 to 0.18 | 0.251 | 0.807 |

β is the change in the outcome per 1 SD better auditory performance: PALTEA z-score units (higher = better) in Great Minds, adjusted for age, sex and education; task score points (4MT, /15; SMT, /12) in PREVENT, adjusted for age and sex. Ordinary least-squares regression with HC3 robust standard errors. q, Benjamini–Hochberg FDR-corrected p-value across the five auditory predictors within each outcome. COGNITO results are shown in Fig 4 and S1 Table. 4MT, Four Mountains Task; SMT, Supermarket Task; other abbreviations as in Table 1.

In PREVENT, no auditory measure was associated with 4MT or SMT scores after age and sex adjustment and FDR correction (Table 3, Fig 3B and 3C). Before correction, better WM-F (β=0.82 points per SD, 95% CI 0.16 to 1.47, p=0.014, q=0.071) and WM-AM (β=0.73, 95% CI 0.06 to 1.39, p=0.032, q=0.081) precision were associated with higher 4MT scores. In unadjusted analyses, 4MT scores correlated with SIB (r=0.35, n=76), WM-F (r=0.33, n=75) and WM-AM (r=0.32, n=75; all q=0.007), indicating that these associations were largely explained by age and sex (S1 Table).

No auditory measure was associated with any COGNITO outcome after FDR correction (n=73– 74; Fig 4, S1 Table). The strongest uncorrected associations were between WM-AM and immediate name recall (β=−0.36, 95% CI −0.67 to −0.04, p=0.028, q=0.14) and between GMSI and semantic fluency (β=0.75, 95% CI 0.01 to 1.48, p=0.047, q=0.20).

**Fig 4.**
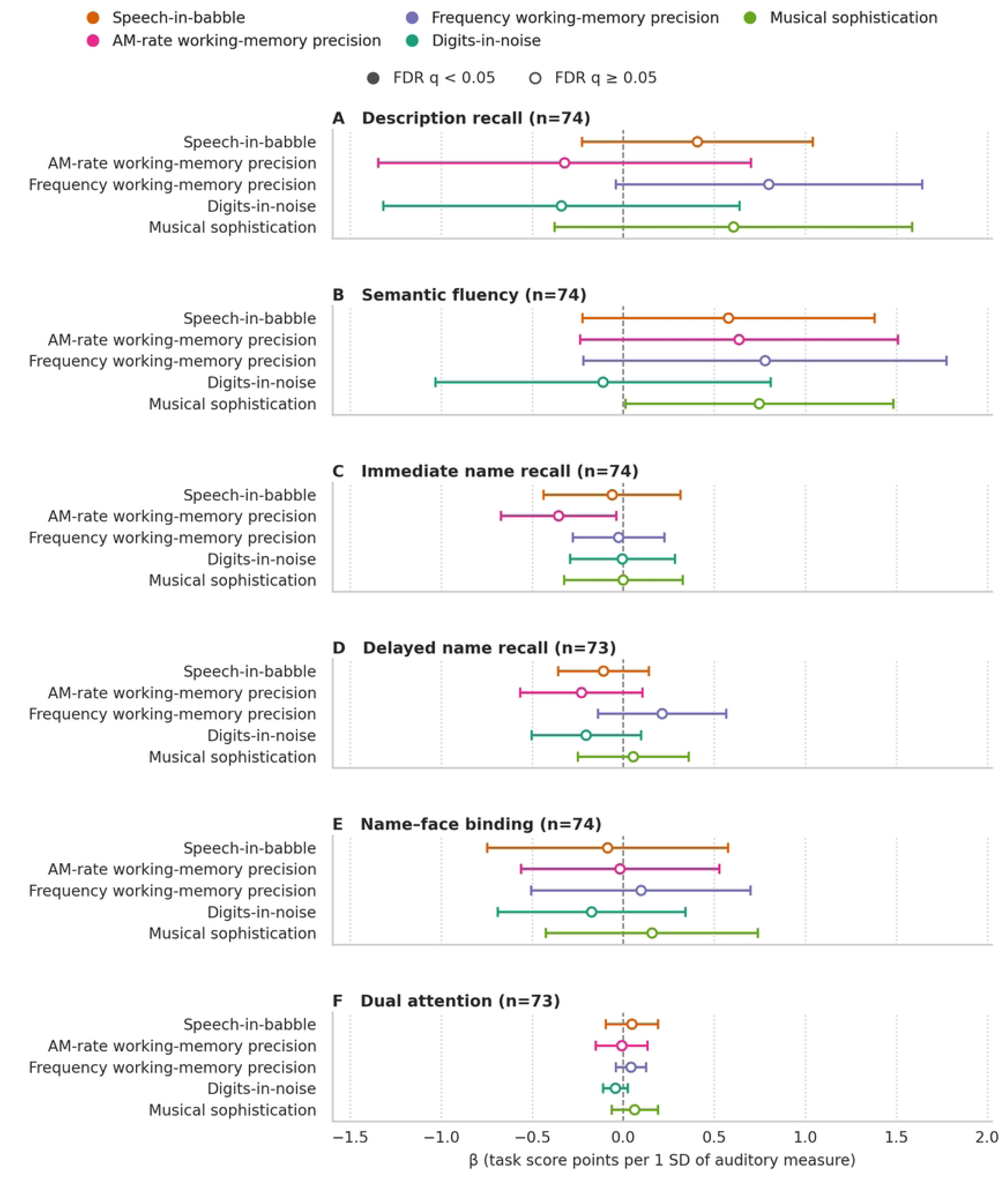
Auditory predictors of COGNITO scores in PREVENT. Points show regression coefficients (β, task score points) per 1 SD better auditory performance, with 95% confidence intervals, for (A) description recall, (B) semantic fluency, (C) immediate name recall, (D) delayed name recall, (E) name–face binding and (F) dual-task attention (visit 1; n=73–74), adjusted for age and sex. Ordinary least-squares regression with HC3 robust standard errors. No estimate reached FDR q < 0.05 within outcome.

## Discussion

Across two cohorts spanning mid-life and later life, APOE ε4 carrier status was not associated with remote auditory task performance, whereas in the older Great Minds cohort better speech-in-babble performance and better working-memory precision for amplitude-modulation rate were associated with better paired-associate learning. In this sample, therefore, variation in remote auditory performance followed cognitive variation more closely than APOE ε4 genotype. The absence of an association between APOE ε4 and auditory measures is consistent with the largest and most directly comparable external study. In the Canadian Longitudinal Study on Aging, APOE ε4 allele count was not associated with pure-tone hearing thresholds, visual acuity or cognition, either cross-sectionally or over three years of follow-up, and the authors concluded that ε4 is unlikely to be a common cause of sensory–cognitive associations [22]. Our findings are consistent with this conclusion for higher-level behavioural auditory measures, namely speech-in-noise perception and auditory working-memory precision, in two cohorts of differing age and risk profile.

The wider literature is mixed. In the population-based Leiden 85-plus Study, APOE ε4 showed a gene-dose relationship with worse pure-tone hearing and a roughly two-fold higher risk of hearing impairment, and a clinical study of 1,162 participants reported an age-dependent, biphasic ε4 effect in which carriers had better hearing at younger ages but greater deterioration with age [23,24]. Several factors may reconcile these findings with ours. First, age and disease stage differ: positive audiometric effects are clearest in the very old (the Leiden participants were all aged 85), whereas our cohorts had mean ages of about 57 (PREVENT) and 71 (Great Minds) and were cognitively healthy. This fits the broader cognitive literature, in which APOE ε4 effects on memory and global cognition are small and become larger with increasing age [7,25]. Under the antagonistic-pleiotropy account, ε4 may even be neutral or advantageous earlier in life and become a liability as amyloid and vascular burden accumulate [26]; a mid-life cohort such as PREVENT may therefore be too early in the trajectory for a behavioural ε4 effect to be detectable. Second, the outcomes differ: most positive reports concern peripheral pure-tone sensitivity, whereas our battery indexes central and cognitive aspects of listening, which need not share the same genetic influences. Third, our samples are research-engaged, cognitively healthy volunteers, which restricts variance and excludes prodromal individuals in whom ε4 effects are most visible. Finally, small ε4-related differences in auditory processing may exist below the resolution of remote behavioural testing.

The auditory–cognition associations we observed are consistent with an established literature. The association of speech-in-babble and amplitude-modulation-rate precision with PALTEA in Great Minds accords with meta-analytic evidence that speech-in-noise perception is reliably, if modestly, associated with cognition in adults [27], and with our previous laboratory finding that short-term memory for sound features predicts both speech-in-noise performance and cognitive screening scores [3]. It is also consistent with prospective evidence that poorer central auditory processing, measured by dichotic listening, predicts incident dementia independently of peripheral hearing [28], and with frameworks linking hearing and dementia through shared central mechanisms [29]. Speech-in-noise and temporal auditory tasks draw on perceptual segregation, sustained attention and auditory short-term memory, processes that overlap with medial-temporal and fronto-temporal systems supporting episodic memory. This provides a plausible basis for auditory performance as a functional correlate of cognition rather than a direct readout of genetic risk.

Auditory–cognition associations were detectable in Great Minds but not in PREVENT. Great Minds is older, with greater sensory and cognitive variability, and its PALTEA analyses had more than twice the sample size of the PREVENT models. In PREVENT, unadjusted correlations between 4MT and auditory measures were attenuated after age and sex adjustment, and COGNITO associations did not survive correction. PREVENT therefore showed the same absence of APOE ε4–auditory associations, but provided no robust evidence of auditory–cognition associations.

Practically, these results support remote speech-in-noise testing as a low-burden, inexpensive and potentially scalable measure of auditory and cognitive function that requires longitudinal validation. Because these measures were not associated with APOE ε4 status in our samples, they should not be interpreted as behavioural proxies for ε4-mediated Alzheimer’s disease risk. A strength of this study is the use of two independent cohorts at different points on the dementia-risk continuum, which allowed the APOE ε4–auditory question to be examined in both an older register and a mid-life prevention cohort. Both cohorts completed the same previously validated online auditory battery with headphone quality control, which reduces measurement differences between cohorts.

Several limitations constrain these conclusions. The absence of APOE ε4 associations should not be read as evidence of no effect: with 40 carriers in Great Minds and fewer in PREVENT, the analyses could detect only moderate-to-large group differences, and the confidence intervals are compatible with small effects in either direction. The cross-sectional design precludes causal inference and cannot detect ε4 effects that emerge only over time, an important caveat given that ε4 effects on cognition are largely an older-age phenomenon. The cohorts were assessed with different cognitive instruments (PALTEA versus 4MT, SMT and COGNITO), so cognitive findings could not be pooled and cross-cohort comparison is qualitative. PREVENT spatial-memory and COGNITO models were smaller (n=68–74) than the Great Minds PALTEA models (n=162–164). The remote auditory assessment was not aligned to the date of each PREVENT clinic visit; 4MT and SMT were taken from visit 2 and COGNITO from visit 1, so variation in the interval between auditory and cognitive assessment may have attenuated associations. The age difference that makes the cohorts complementary also means that cohort and age are partly confounded. Both cohorts consist of research-engaged, UK-based volunteers who are cognitively healthier and more educated than the general population, which limits generalisability and, by compressing variance, may attenuate the associations under study. Finally, remote testing carries device and environmental variability that, despite quality control, adds noise and may obscure small effects [8].

Taken together, these findings support remote auditory testing as a scalable behavioural approach to cognitive phenotyping in ageing and dementia-risk research, while indicating that it should not be interpreted as a proxy for APOE ε4-mediated risk. Longitudinal studies are needed to determine whether baseline speech-in-noise and temporal auditory measures predict subsequent cognitive decline or biomarker change, and whether APOE ε4 effects emerge as these cohorts age.

## Conclusions

Across an older research-register cohort and a mid-life prevention cohort, APOE ε4 carrier status was not associated with remote auditory task performance, whereas in the older cohort speech-in-babble performance and auditory memory for temporal fluctuations were associated with memory performance. Remote speech-in-noise testing may offer a practical, scalable adjunct for cognitive characterisation in ageing cohorts.

## Data Availability

The data underlying this study are third-party data that cannot be shared publicly because they contain personal and health information and are governed by participant consent and data access agreements. Great Minds data are available to bona fide researchers on application to Dementias Platform UK (DPUK) via the DPUK Data Portal (https://portal.dementiasplatform.uk/). PREVENT Dementia programme data are available on application to the PREVENT Steering Committee via the DPUK Data Portal or the Alzheimer’s Disease Data Initiative (https://www.alzheimersdata.org/).

## Acknowledgments

We thank the participants and research teams of Great Minds and the PREVENT Dementia programme, and Dementias Platform UK (DPUK) for data access. PREVENT clinical, genetic and cognitive data were provided by the PREVENT data custodians. ML was supported by a Medical Research Council (MRC) Clinical Research Training Fellowship (MR/V006568/1).

## Supporting information

**S1 Checklist. STROBE checklist.** Completed STROBE checklist for cross-sectional studies.

**S1 Table. Full regression and correlation results.** All APOE ε4–auditory models; all auditory– cognition models for PALTEA, Four Mountains and Supermarket (visit 2 primary and baseline sensitivity) and COGNITO outcomes; and unadjusted PREVENT auditory–spatial-memory correlations.

## Notes

### Competing Interest Statement

The authors have declared no competing interest.

### Author Declarations

This study was approved by the Newcastle University Faculty of Medical Sciences Ethics Committee (2682/41403).

